# COVID-19 case-fatality rate and demographic and socioeconomic influencers: a worldwide spatial regression analysis based on country-level data

**DOI:** 10.1101/2020.07.31.20165811

**Authors:** Yang Cao, Ayako Hiyoshi, Scott Montgomery

## Abstract

We used the COVID-19 dataset obtained from the Our World in Data website and investigated the associations between COVID-19 CFR and nine country-level indices of 209 countries/territories using the Matern correlation regression model. Spatial dependence among the data was controlled using the latitude and longitude of the centroid of the countries/territories. Stratified analyses were conducted by economic level and COVID-19 testing policy. The average of country/territory-specific COVID-19 CFR is about 2-3% worldwide, which is higher than previously reported at 0.7-1.3%. Statistically significant associations were observed between COVID-19 CFR and population size and proportion of female smokers. The open testing policies are associated with decreased CFR. Strictness of anti-COVID-19 measures was not statistically significantly associated with CFR overall, but the higher stringency index was associated with higher CFR in higher income countries with active testing policies. The statistically significant association between population size and COVID-19 CRF suggests the healthcare strain and lower treatment efficiency in countries with large populations. The observed association between smoking in females and COVID-19 CFR might be due to that the proportion of female smokers reflected broadly income level of a country. When testing is warranted and healthcare resources are sufficient, strict quarantine and/or lockdown measures might result in excess deaths in underprivileged populations.

## Introduction

The pandemic caused by the severe acute respiratory syndrome coronavirus 2 (SARS-COV2 Virus/COVID-19), which was initially reported in Wuhan, a city in the Hubei province in China in December 2019, has become a major global health concern (Margallo II et al. 2020). Poor outcomes in those with COVID□19 infections correlate with clinical and laboratory features of cytokine storm syndrome, an exaggerated systemic inflammatory phenomenon due to over-production of proinflammatory cytokines by the immune system that results in diffuse inflammatory lung disease and acute respiratory distress syndrome (Henderson et al. 2020). It may be complicated by sepsis, respiratory failure, acute respiratory distress syndrome (ARDS), and subsequent multi-organ failure (Zaim et al. 2020). Although COVID-19 related deaths are not clearly defined in the international reports available so far, many governments are warning people to be particularly stringent in following the recommended prevention measures because COVID-19 may results in severe conditions that need critical care including ventilation or death (Jordan et al. 2020). Till the end of June, 2020, the pandemic has resulted in over 10 million confirmed cases and 500 thousand deaths worldwide (Worldometer 2020). COVID-19 related fatality rates are difficult to assess with certainty, but according to the estimates based on the data from China, the United Kingdom, Italy, and the Diamond Princess cruise ship, the overall death rate from the confirmed COVID-19 cases is around 0.7-1.3%, with a sharply rising from less than 0.002% in children aged 9 years or younger to 8% in people aged over 80, which is much greater than seasonal influenza at about 0.1% (Baud et al. 2020; Jordan et al. 2020; Mahase 2020; Onder et al. 2020; Rajgor et al. 2020; Russell et al. 2020).

During the COVID-19 pandemic, numerous studies on the global public health emergency have covered a significant range of disciplines including medicine, mathematics and social sciences. The spatial spread is one of the most important properties of COVID-19, a characteristic which mainly depends on the epidemic mechanism, human mobility, and control strategy (Gross et al. 2020). Spatial statistical methods are frequently used to uncover relationships between spatiotemporal patterns of infectious diseases and host or environmental characteristics (Lawson 2013), generate detailed maps to visualize the distribution of the diseases’ morbidity or mortality (Palk and Blower 2018; Zulu et al. 2014), and identify hotspots, clusters, and potential risk factors (Anselin et al. 2006; Kulldorff and Nagarwalla 1995; Pfeiffer et al. 2008; Sasaki et al. 2008).

Clinical risk factors for mortality of adult patients with COVID-19 have been investigated in numerous studies, and the identified factors include older age (Brooke and Jackson 2020), male sex (Jin et al. 2020), higher sequential organ failure assessment score (Zhou et al. 2020), obesity (Klang et al. 2020; Zhang et al. 2020), preexisting concurrent diabetes (Guo et al. 2020), cardiovascular, cerebrovascular (Du et al. 2020), and kidney diseases (Cheng et al. 2020b), and macro-economic and environmental risk factors such as socioeconomic deprivation (Bibbins-Domingo 2020), air pollution (Ogen 2020; Wu et al. 2020) and diurnal temperature variation (Ma et al. 2020), etc. However, there is a lack of published studies on the effects of demographic and socioeconomic factors on COVID-10 case-fatality rates at the population level. It is an important issue for governments and regional or international non-governmental organizations to identify country characteristics that are associated with high case fatality rate (CRF) and help developing prevention and intervention measures to fight against this global public health crisis.

Using the publicly available data from the non-governmental organization Our World in Data (Our World in Data 2020a), we aimed to investigate the relationship of key country level demographic and socioeconomic indices and COVID-19 case-fatality, and explore factors associated with CFR, which may indicate treatment efficiency and strain in healthcare resources, while controlling for the spatial dependence of the data collected at different locations.

The data used in the study are freely available in the open source for research. There are no individual data available in the dataset. The owner of the data gives the permission to use, distribute, and reproduce the data in any medium. Therefore, no private and confidential information could be disclosed in the study, and ethical approval is not applicable. In the study, estimates of health indicators at the global level were reported according to the GATHER statement (Supplemental material 1) (Stevens et al. 2016).

## Results

### Descriptive characteristics of the variables

In total, 209 countries and territories and 17 variables (including latitude and longitude) were included in the study. The descriptive statistics of the variables are shown in Tables 1. The pairwise relationships of the variables are shown in Figure 1. High multicollinearity was found between number of confirmed cases and population size (pairwise Pearson’s r = 0.76, P < 0.001, Figure 1), and gross domestic product (GDP) per capita and life expectancy, median age, proportion of aged 65 years or older, proportion of extreme poverty, and hospital beds per 1000 people (all pairwise Pearson’s r greater than or approximate to 0.70, and P < 0.001, Figure 1; multiple correlation coefficient = 0.92, P < 0.001). Therefore, median age, proportion of aged 65 or older, and hospital beds per 1000 people were excluded in later regression analysis. Although proportion of extreme poverty is an interesting factor to be investigated, it is highly correlated with GDP per capita (r = −0.83) and the latter is available in more countries, therefore proportion of extreme poverty was also excluded from analysis.

**Table 1.** Descriptive statistics of the variables

|  | N | Mean | SD | Median | Min | Max | IQR |
| --- | --- | --- | --- | --- | --- | --- | --- |
| Covid-19 CFR (%) | 209 | 3.31 | 3.79 | 2.19 | 0.00 | 26.94 | 3.75 |
| Covid-19 CDR (per 1,000,000 person months) | 209 | 13.98 | 28.75 | 3.31 | 0.00 | 205.84 | 10.94 |
| Confirmed cases (log) | 209 | 7.75 | 2.84 | 7.71 | 1.10 | 14.78 | 4.13 |
| Population size (log) | 209 | 15.28 | 2.52 | 15.74 | 6.70 | 21.09 | 3.29 |
| GDP per capita (log) | 182 | 9.28 | 1.21 | 9.48 | 6.49 | 11.67 | 1.84 |
| Population density (log) | 198 | 4.42 | 1.52 | 4.47 | -1.99 | 9.87 | 1.74 |
| CVD death rate (log) | 185 | 5.44 | 0.46 | 5.49 | 4.37 | 6.59 | 0.65 |
| Diabetes prevalence (log) | 192 | 1.94 | 0.56 | 1.96 | -0.01 | 3.15 | 0.67 |
| Stringency index | 172 | 44.22 | 9.16 | 45.46 | 7.68 | 65.20 | 11.16 |
| Life expectancy | 206 | 73.47 | 7.54 | 75.07 | 53.28 | 86.75 | 9.91 |
| Median age | 185 | 30.55 | 9.10 | 29.90 | 15.10 | 48.20 | 16.50 |
| Age 65 years or older (log) | 182 | 1.91 | 0.74 | 1.90 | 0.13 | 3.30 | 1.42 |
| Proportion of extreme poverty (log) | 121 | 1.08 | 2.03 | 0.79 | -2.30 | 4.35 | 4.57 |
| Proportion of female smoker (log) | 140 | 1.63 | 1.40 | 1.80 | -2.30 | 3.78 | 2.31 |
| Proportion of male smoker | 138 | 32.62 | 13.71 | 31.80 | 7.70 | 78.10 | 19.90 |
| Hospital beds per 1000 people (log) | 164 | 0.77 | 0.88 | 0.86 | -2.30 | 2.62 | 1.11 |

SD, standard deviation; IQR, interquartile range; GDP, gross domestic product; CVD, cardiovascular disease.

**Figure 1.**
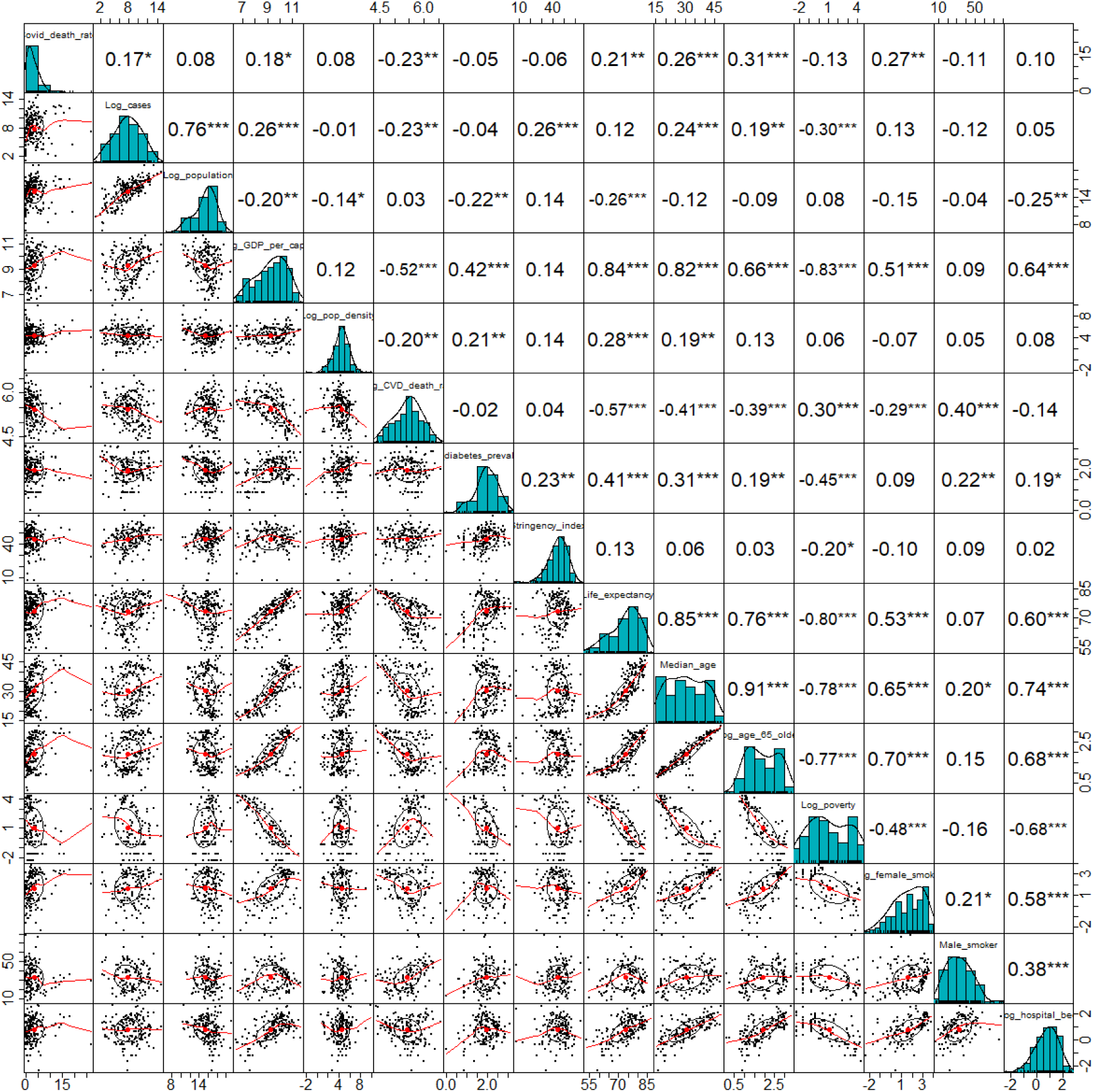
Pairwise scatter plots and Pearson’s correlation coefficients of the variables ***, P<0.001; **, P<0.01; *, P<0.05

### Worldwide COVID-19 CFR distribution

Distribution of COVID-19 case fatality rates of the 209 countries/territories is show in Figure 2. The mean and median CFR worldwide are 3.31% and 2.19%, respectively (Table 1), with the highest rates found in Yeman (27%), West and North Europe (14-19%), and North America (9-12%, Figure 2).

**Figure 2.**
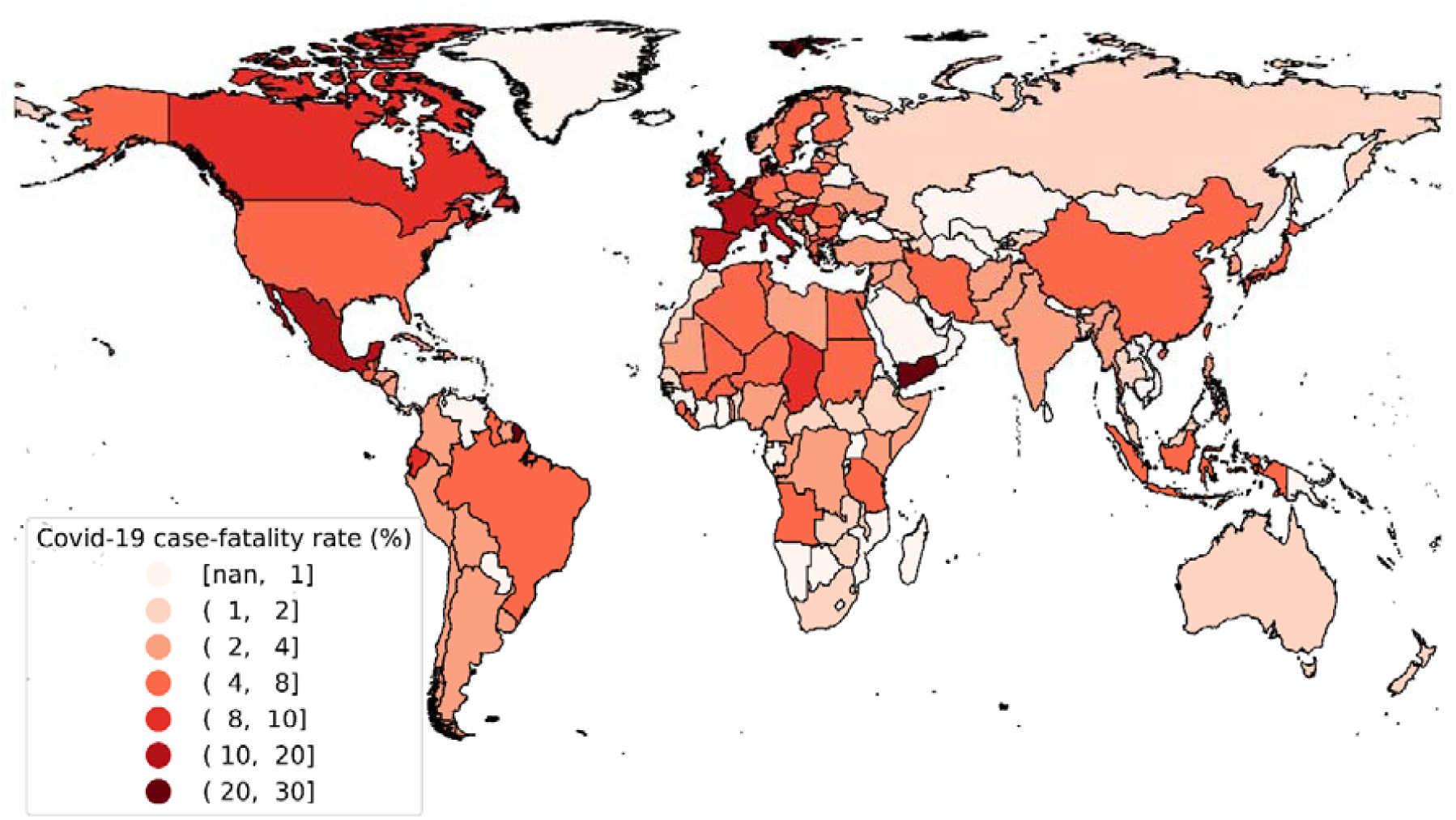
Global COVID-19 case-fatality rates (%) nan: no data available

### Spatial autocorrelation of the COVID-19 CFR

Statistically significant spatial autocorrelation was found among the countries/territories’ COVID-19 case-fatality rates. The residuals from the common (non-spatial) multivariate linear regression models show apparent spatial dependence around the countries/territories with high fatality (Figure 3). The P-value from the Moran’s I test for the spatial autocorrelation of the residuals is 2.32×10^−5^.

**Figure 3.**
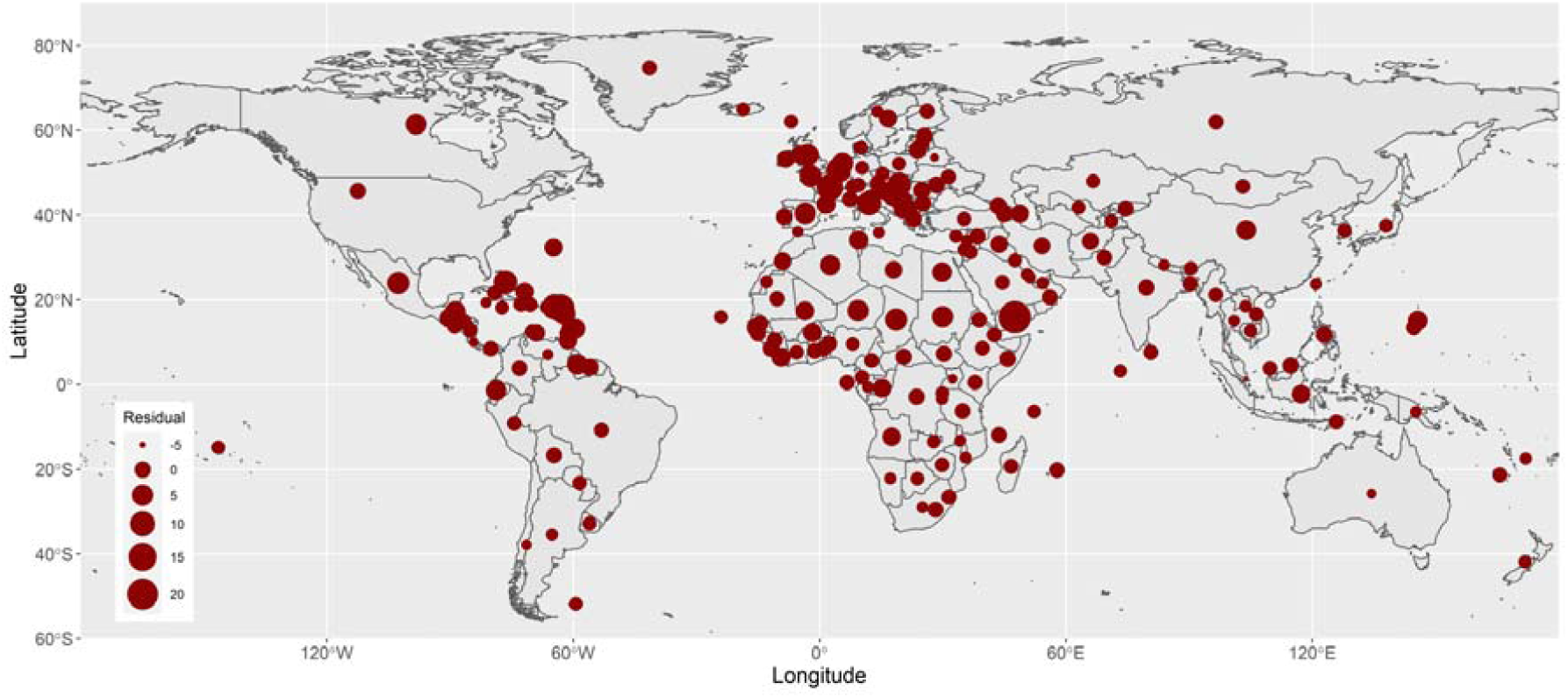
Residuals of the common (non-spatial) multivariate linear regression

The estimated spatial autocorrelation coefficient of COVID-19 case-fatality rates between two locations against their distance is shown in Figure 4, with a strength parameter *ν* of 2.48 and a decay parameter *ρ* of 0.12. Basically, locations more than 20 degrees (in longitude or latitude) away have an autocorrelation coefficient below 0.5 (Figure 4).

**Figure 4.**
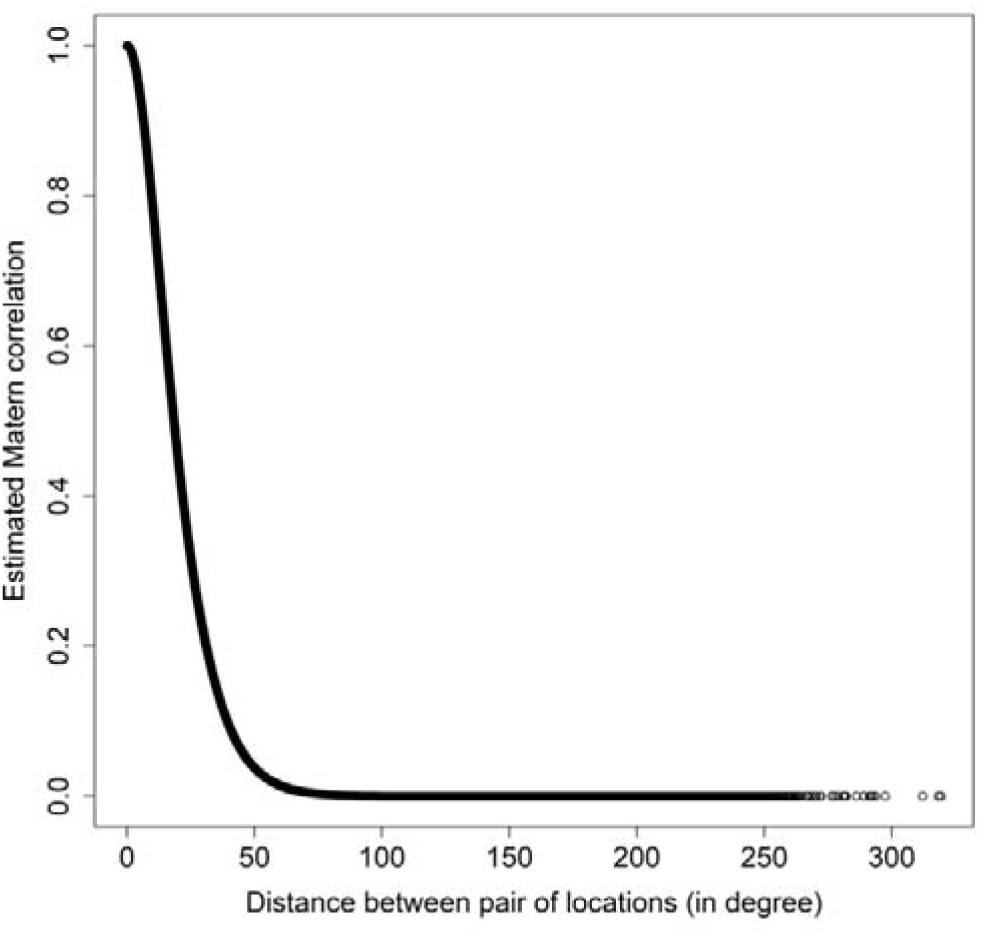
Strength and decay of the spatial autocorrelation between pair of locations

### Associations of demographic and socioeconomic variables with COVID-19 CFR

Overall, after controlling for the spatial dependence, the statistically significant variables associated with COVID-19 case-fatality rate are population size and proportion of female smokers in a country/region (Table 2). The multivariate adjusted results indicate that, approximately, a doubling in size of population is associated with 0.48 (95% confidence interval (CI): 0.25, 0.70) percent increase in COVID-19 case-fatality rate, and a doubling in proportion of female smokers is associated with 0.55 (95% CI: 0.09, 1.02) percent increase in in COVID-19 case-fatality rate. Open public testing policy is associated with decreased case-fatality rate (Beta = 2.23, 95% CI: −4.25, −0.21) percent compared to no testing policy. Although not statically significant, stringency index showed the smallest coefficient with COVID-19 case-fatality rate (Beta = −0.02, 95% CI: −0.08, 0.03) among all the investigated variables (Table 2).

**Table 2.**
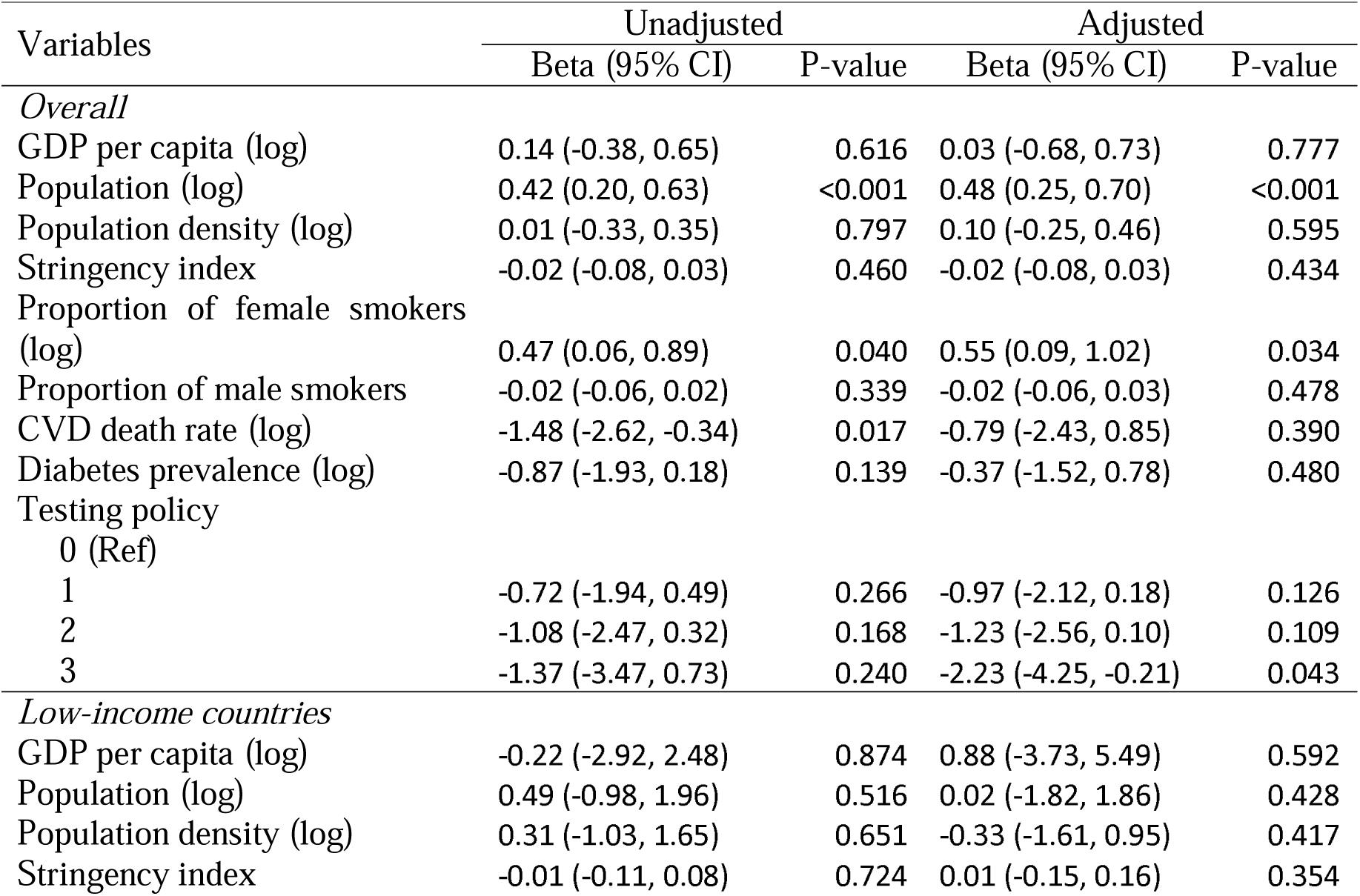
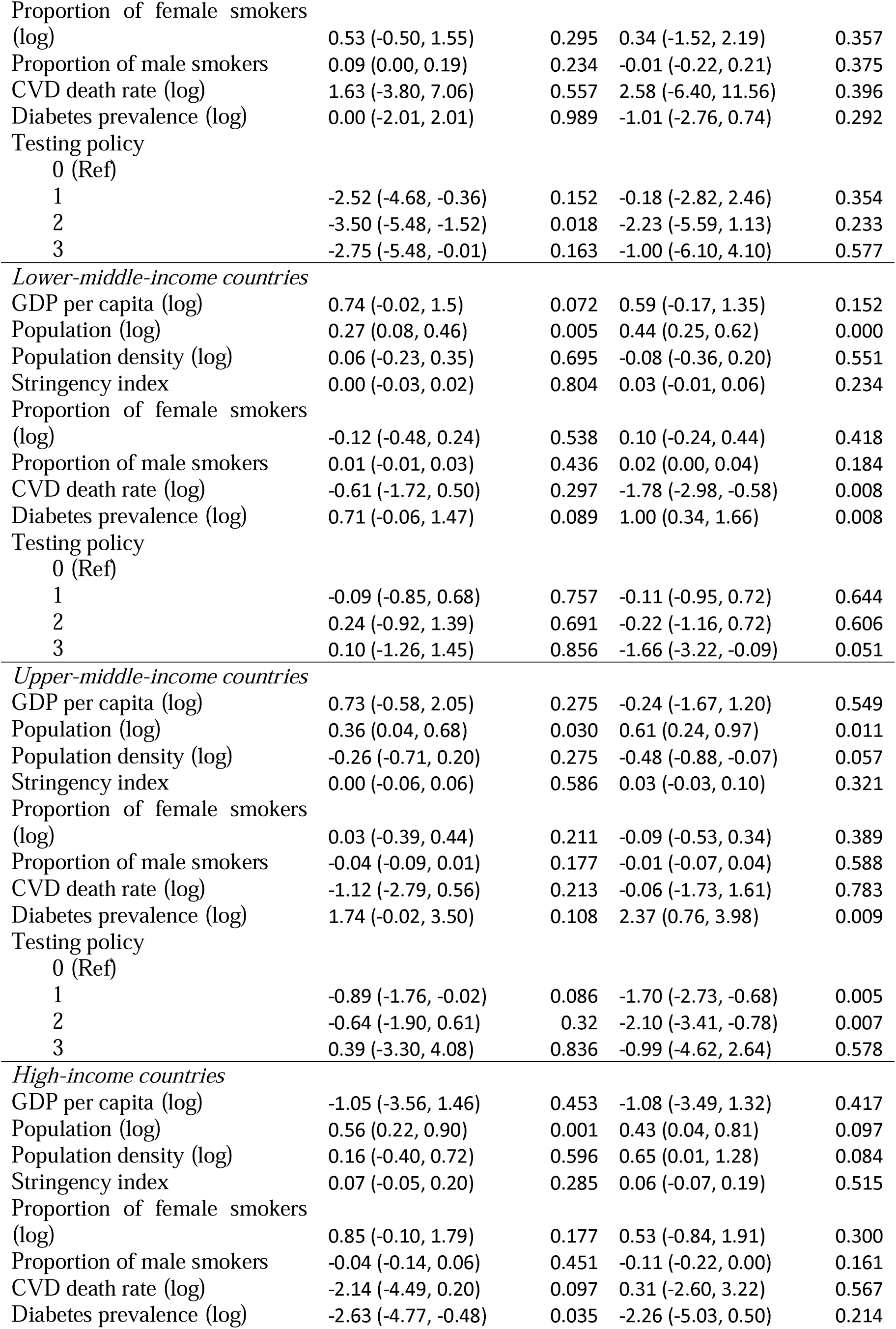
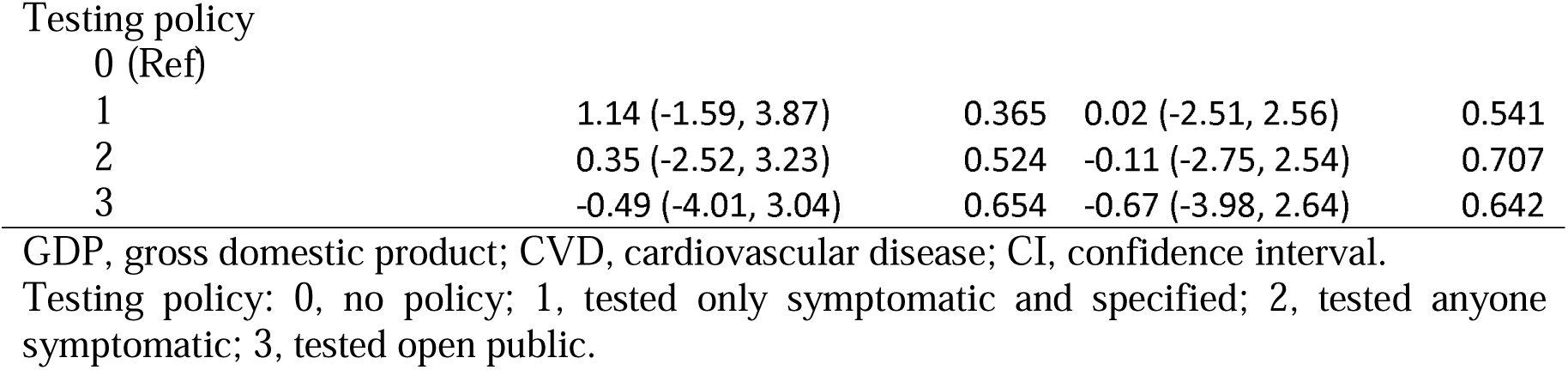
Estimation for variables’ regression coefficients (Beta) for CFR

| Variables | Unadjusted |  | Adjusted |  |
| --- | --- | --- | --- | --- |
|  | Beta (95% CI) | P-value | Beta (95% CI) | P-value |
| *Overall* |  |  |  |  |
| GDP per capita (log) | 0.14 (-0.38, 0.65) | 0.616 | 0.03 (-0.68, 0.73) | 0.777 |
| Population (log) | 0.42 (0.20, 0.63) | <0.001 | 0.48 (0.25, 0.70) | <0.001 |
| Population density (log) | 0.01 (-0.33, 0.35) | 0.797 | 0.10 (-0.25, 0.46) | 0.595 |
| Stringency index | -0.02 (-0.08, 0.03) | 0.460 | -0.02 (-0.08, 0.03) | 0.434 |
| Proportion of female smokers (log) | 0.47 (0.06, 0.89) | 0.040 | 0.55 (0.09, 1.02) | 0.034 |
| Proportion of male smokers | -0.02 (-0.06, 0.02) | 0.339 | -0.02 (-0.06, 0.03) | 0.478 |
| CVD death rate (log) | -1.48 (-2.62, -0.34) | 0.017 | -0.79 (-2.43, 0.85) | 0.390 |
| Diabetes prevalence (log) | -0.87 (-1.93, 0.18) | 0.139 | -0.37 (-1.52, 0.78) | 0.480 |
| Testing policy |  |  |  |  |
| 0 (Ref) |  |  |  |  |
| 1 | -0.72 (-1.94, 0.49) | 0.266 | -0.97 (-2.12, 0.18) | 0.126 |
| 2 | -1.08 (-2.47, 0.32) | 0.168 | -1.23 (-2.56, 0.10) | 0.109 |
| 3 | -1.37 (-3.47, 0.73) | 0.240 | -2.23 (-4.25, -0.21) | 0.043 |
| *Low-income countries* |  |  |  |  |
| GDP per capita (log) | -0.22 (-2.92, 2.48) | 0.874 | 0.88 (-3.73, 5.49) | 0.592 |
| Population (log) | 0.49 (-0.98, 1.96) | 0.516 | 0.02 (-1.82, 1.86) | 0.428 |
| Population density (log) | 0.31 (-1.03, 1.65) | 0.651 | -0.33 (-1.61, 0.95) | 0.417 |
| Stringency index | -0.01 (-0.11, 0.08) | 0.724 | 0.01 (-0.15, 0.16) | 0.354 |
| Proportion of female smokers (log) | 0.53 (-0.50, 1.55) | 0.295 | 0.34 (-1.52, 2.19) | 0.357 |
| Proportion of male smokers | 0.09 (0.00, 0.19) | 0.234 | -0.01 (-0.22, 0.21) | 0.375 |
| CVD death rate (log) | 1.63 (-3.80, 7.06) | 0.557 | 2.58 (-6.40, 11.56) | 0.396 |
| Diabetes prevalence (log) | 0.00 (-2.01, 2.01) | 0.989 | -1.01 (-2.76, 0.74) | 0.292 |
| Testing policy |  |  |  |  |
| 0 (Ref) |  |  |  |  |
| 1 | -2.52 (-4.68, -0.36) | 0.152 | -0.18 (-2.82, 2.46) | 0.354 |
| 2 | -3.50 (-5.48, -1.52) | 0.018 | -2.23 (-5.59, 1.13) | 0.233 |
| 3 | -2.75 (-5.48, -0.01) | 0.163 | -1.00 (-6.10, 4.10) | 0.577 |
| *Lower-middle-income countries* |  |  |  |  |
| GDP per capita (log) | 0.74 (-0.02, 1.5) | 0.072 | 0.59 (-0.17, 1.35) | 0.152 |
| Population (log) | 0.27 (0.08, 0.46) | 0.005 | 0.44 (0.25, 0.62) | 0.000 |
| Population density (log) | 0.06 (-0.23, 0.35) | 0.695 | -0.08 (-0.36, 0.20) | 0.551 |
| Stringency index | 0.00 (-0.03, 0.02) | 0.804 | 0.03 (-0.01, 0.06) | 0.234 |
| Proportion of female smokers (log) | -0.12 (-0.48, 0.24) | 0.538 | 0.10 (-0.24, 0.44) | 0.418 |
| Proportion of male smokers | 0.01 (-0.01, 0.03) | 0.436 | 0.02 (0.00, 0.04) | 0.184 |
| CVD death rate (log) | -0.61 (-1.72, 0.50) | 0.297 | -1.78 (-2.98, -0.58) | 0.008 |
| Diabetes prevalence (log) | 0.71 (-0.06, 1.47) | 0.089 | 1.00 (0.34, 1.66) | 0.008 |
| Testing policy |  |  |  |  |
| 0 (Ref) |  |  |  |  |
| 1 | -0.09 (-0.85, 0.68) | 0.757 | -0.11 (-0.95, 0.72) | 0.644 |
| 2 | 0.24 (-0.92, 1.39) | 0.691 | -0.22 (-1.16, 0.72) | 0.606 |
| 3 | 0.10 (-1.26, 1.45) | 0.856 | -1.66 (-3.22, -0.09) | 0.051 |
| *Upper-middle-income countries* |  |  |  |  |
| GDP per capita (log) | 0.73 (-0.58, 2.05) | 0.275 | -0.24 (-1.67, 1.20) | 0.549 |
| Population (log) | 0.36 (0.04, 0.68) | 0.030 | 0.61 (0.24, 0.97) | 0.011 |
| Population density (log) | -0.26 (-0.71, 0.20) | 0.275 | -0.48 (-0.88, -0.07) | 0.057 |
| Stringency index | 0.00 (-0.06, 0.06) | 0.586 | 0.03 (-0.03, 0.10) | 0.321 |
| Proportion of female smokers (log) | 0.03 (-0.39, 0.44) | 0.211 | -0.09 (-0.53, 0.34) | 0.389 |
| Proportion of male smokers | -0.04 (-0.09, 0.01) | 0.177 | -0.01 (-0.07, 0.04) | 0.588 |
| CVD death rate (log) | -1.12 (-2.79, 0.56) | 0.213 | -0.06 (-1.73, 1.61) | 0.783 |
| Diabetes prevalence (log) | 1.74 (-0.02, 3.50) | 0.108 | 2.37 (0.76, 3.98) | 0.009 |
| Testing policy |  |  |  |  |
| 0 (Ref) |  |  |  |  |
| 1 | -0.89 (-1.76, -0.02) | 0.086 | -1.70 (-2.73, -0.68) | 0.005 |
| 2 | -0.64 (-1.90, 0.61) | 0.32 | -2.10 (-3.41, -0.78) | 0.007 |
| 3 | 0.39 (-3.30, 4.08) | 0.836 | -0.99 (-4.62, 2.64) | 0.578 |
| *High-income countries* |  |  |  |  |
| GDP per capita (log) | -1.05 (-3.56, 1.46) | 0.453 | -1.08 (-3.49, 1.32) | 0.417 |
| Population (log) | 0.56 (0.22, 0.90) | 0.001 | 0.43 (0.04, 0.81) | 0.097 |
| Population density (log) | 0.16 (-0.40, 0.72) | 0.596 | 0.65 (0.01, 1.28) | 0.084 |
| Stringency index | 0.07 (-0.05, 0.20) | 0.285 | 0.06 (-0.07, 0.19) | 0.515 |
| Proportion of female smokers (log) | 0.85 (-0.10, 1.79) | 0.177 | 0.53 (-0.84, 1.91) | 0.300 |
| Proportion of male smokers | -0.04 (-0.14, 0.06) | 0.451 | -0.11 (-0.22, 0.00) | 0.161 |
| CVD death rate (log) | -2.14 (-4.49, 0.20) | 0.097 | 0.31 (-2.60, 3.22) | 0.567 |
| Diabetes prevalence (log) | -2.63 (-4.77, -0.48) | 0.035 | -2.26 (-5.03, 0.50) | 0.214 |
| Testing policy |  |  |  |  |
| 0 (Ref) |  |  |  |  |
| 1 | 1.14 (-1.59, 3.87) | 0.365 | 0.02 (-2.51, 2.56) | 0.541 |
| 2 | 0.35 (-2.52, 3.23) | 0.524 | -0.11 (-2.75, 2.54) | 0.707 |
| 3 | -0.49 (-4.01, 3.04) | 0.654 | -0.67 (-3.98, 2.64) | 0.642 |

GDP, gross domestic product; CVD, cardiovascular disease; CI, confidence interval.
Testing policy: 0, no policy; 1, tested only symptomatic and specified; 2, tested anyone symptomatic; 3, tested open public.

The estimated contour of COVID-19 case-fatality rates worldwide is shown in Figure 5. The areas with the higher risks are mainly around North America and West Europe.

**Figure 5.**
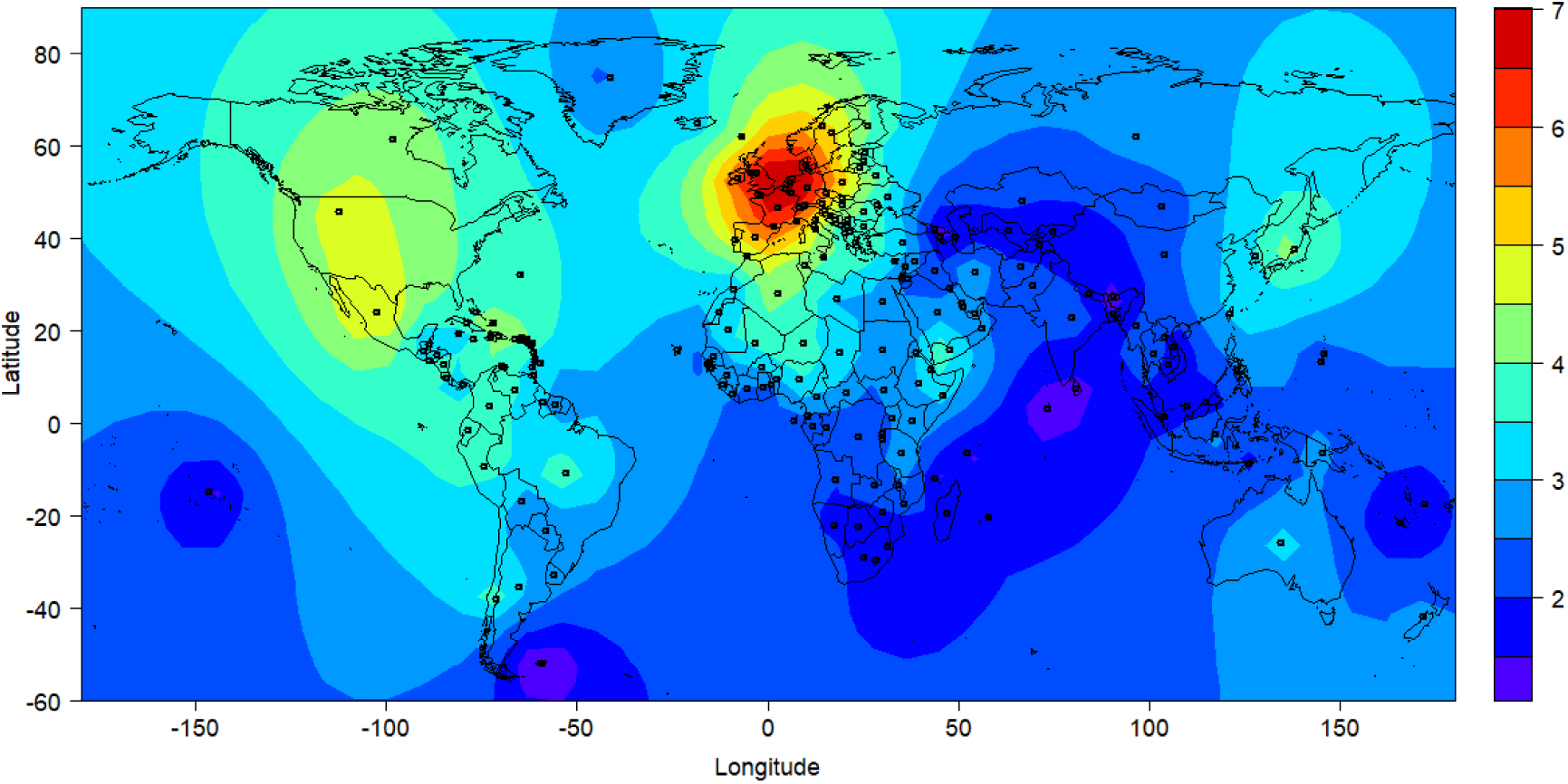
Contour plot of estimated COVID-19 case-fatality rate (%)

The subgroup analysis by economic level indicates that population size, CVD death rate, diabetes prevalence, and open public testing policy; population size, population density, diabetes prevalence, and testing only symptomatic and specified policy and testing anyone symptomatic policy; and population size, and population density are statistically significantly associated with COVID-19 case-fatality rate in lower-middle, upper-middle, and high income countries, respectively (Table 2).

However, the subgroup analysis in upper-middle-income and high-income countries by testing policy indicates that, if testing was ensured (testing policy =2 or 3), stringency index is associated with increased case-fatality rate (Beta = 0.14, 95% CI: 0.01, 0.27) (Table 3). The find suggests an open society policy would be good when testing is ensured. Meanwhile, diabetes prevalence is inversely associated with case-fatality rate.

**Table 3.** Estimation for variables’ regression coefficients (Beta) for CFR by testing policy in upper-middle-income and high-income countries

| Variables | Unadjusted |  | Adjusted |  |
| --- | --- | --- | --- | --- |
|  | Beta (95% CI) | P-value | Beta (95% CI) | P-value |
| *Testing policy=1 (tested only symptomatic & specified)* |  |  |  |  |
| GDP per capita (log) | 0.51 (-1.05, 2.08) | 0.522 | -0.22 (-2.04, 1.61) | 0.805 |
| Population (log) | 0.34 (-0.29, 0.97) | 0.290 | 0.32 (-0.34, 0.99) | 0.393 |
| Population density (log) | 0.49 (-0.15, 1.13) | 0.132 | 0.58 (-0.15, 1.30) | 0.131 |
| Stringency index | -0.07 (-0.22, 0.08) | 0.349 | -0.06 (-0.23, 0.10) | 0.469 |
| Proportion of female smokers (log) | 0.20 (-0.96, 1.36) | 0.667 | 0.13 (-1.28, 1.55) | 0.409 |
| Proportion of male smokers | -0.08 (-0.19, 0.03) | 0.186 | -0.04 (-0.20, 0.11) | 0.555 |
| CVD death rate (log) | -1.69 (-3.91, 0.52) | 0.137 | -0.20 (-3.70, 3.30) | 0.798 |
| Diabetes prevalence (log) | -0.65 (-3.52, 2.22) | 0.659 | -0.82 (-4.37, 2.73) | 0.672 |
| *Testing policy=2 or 3 (tested anyone symptomatic or anyone public)* |  |  |  |  |
| GDP per capita (log) | 0.35 (-1.42, 2.11) | 0.706 | 0.45 (-1.19, 2.09) | 0.590 |
| Population (log) | 0.81 (0.38, 1.23) | 0.000 | 0.49 (0.03, 0.96) | 0.047 |
| Population density (log) | 0.08 (-0.55, 0.7) | 0.812 | 0.02 (-0.61, 0.65) | 0.913 |
| Stringency index | 0.17 (0.05, 0.28) | 0.005 | 0.14 (0.01, 0.27) | 0.044 |
| Proportion of female smokers (log) | 0.70 (-0.22, 1.61) | 0.151 | 0.40 (-0.52, 1.33) | 0.400 |
| Proportion of male smokers | 0.00 (-0.10, 0.10) | 0.890 | 0.00 (-0.11, 0.11) | 0.845 |
| CVD death rate (log) | -2.34 (-4.66, -0.02) | 0.049 | -2.00 (-4.95, 0.94) | 0.189 |
| Diabetes prevalence (log) | -2.45 (-5.21, 0.32) | 0.089 | -3.30 (-5.85, -0.74) | 0.013 |

## Discussion

### Geospatial analysis of the COVID-19 pandemic

The COVID-19 pandemic is still full of unknowns, and many of them have a spatial dimension that lead to understanding the emergency geographically. Analysis of the COVID-19 data requires an interdisciplinary approach, including spatial statistics that may provide solutions to address the spatial issues in the pandemic(Meade 2014). A recently published review summarized studies by May 1, 2020 on geospatial and spatial-statistical analysis of the COVID-19 pandemic. In total 63 studies were categorized into five subjects: spatiotemporal analysis, health and social geography, environmental variables, data mining, and web-based mapping (Franch-Pardo et al. 2020). Although 15 of the studies address the question globally, none of them investigate the association of COVID-19 related deaths with country level demographic and/or socioeconomic factors. From a global health perspective, there is apparent knowledge gap in the research field. Our geospatial analysis fills this gap and help to understand the consequences of COVID-19 and their related factors from a global level, and contribute to the predictive modelling and decision-making to combat the pandemic.

### Population size and density and COVID-19 CFR

Our results indicate that larger population size is the most consistently associated with higher COVID-19 CFR, but population density was not associated with the outcome, controlled for some demographic and socioeconomic variables and spatial dependence worldwide. The association of population size with CFR can be interpreted at least two ways. It could be because larger countries experienced greater number of death and conducted relatively less test compared with countries with smaller population. Alternatively, in larger countries, healthcare might have been strained and resulted in relatively higher number of deaths among confirmed cases than smaller countries (World Health Organization 2020a). We are unable to disentangle the mechanisms of the association. Therefore, it is recommended that the analysis is replicated by a study with more detailed healthcare information on both individual- and country-levels. However, in national studies, higher density has been shown to associate with higher Covid-19 prevalence in Japan (Bassino and Ladmiral 2020), Italy(Sjodin et al. 2020), and Iran (Ahmadi et al. 2020). The lack of the association of COVID-19 CFR with population density globally might be due to the confounding by testing strategies and economic levels. In countries like Germany and South Korea, which took more active testing strategy than, for example, the United Kingdom (UK) where polymerase chain reaction (PCR) test for COVID-19 was only performed among those who were with severe symptoms and hospital admitted at the beginning (Cheng et al. 2020a), CFR naturally show lower values. There are weak but statistically significant negative associations between population size and population density (r = −0.15, p = 0.042) and GDP per capita (r = −0.20, p = 0.006). In order to minimize the confounding, we conducted stratified analysis by economic level (Table 2) and testing policy (only within upper-middle-income and high-income countries, Table 3). The results indicate that in high-income countries, higher population density was associated with increased COVID-19 CFR (Table 2). Furthermore, we conducted a sensitivity analysis (Appendix) for CDR and the results were similar (Table S1 and S2). The results suggests that, globally, healthcare strain should be firstly relieved and treatment efficiency should be improved in countries with large populations.

### Economic level and COVID-19 CFR

In our analysis, high COVID-19 CFR was found mainly around North America and West Europe (Figure 1). One of the possible reasons might be that these countries counted COVID-19 deaths by including those who died with it, not only from it (Onder et al. 2020; Schellekens and Sourrouille 2020). Determination of COVID-19 deaths also differed by country. Some countries recorded a COVID-19 death as any death once the patient became a confirmed case, even the death happened after 2 months possibly by other reasons (such as an accident), while in some other countries, a COVID-19 death was recorded as the death occurring within a certain time period (ranging from 2 to 8 weeks) after COVID-19 symptom onset (Our World in Data 2020b). Furthermore, the extent that the counting covered home, institutions, and hospitals in high-income countries is different with that in low-income countries (Schellekens and Sourrouille 2020).

It has been reported in earlier studies that CFR was more favorable in low-income countries (Cash and Patel 2020; Schellekens and Sourrouille 2020). There are three possibilities to explain this unusual pattern: it may be because of younger population, poor data quality, or it was still the early stage (till we wrote this paper) of COVID-19 infection (Schellekens and Sourrouille 2020). There is a tight relationship between income level of a country and demographic structure. For example, many of African countries were classified as low-income, with median age of 20 years, and 61% of population of them is 24 years or younger, and merely 3% is equal to or older than 65 years in 2015 (United Nations 2019). It has been shown that younger age is associated with lower likelihood of severe COVID-19 (Jordan et al. 2020; Li et al. 2020; Team 2020). However, age-related explanation may not be satisfactory given other factors that are usually associated with higher spread and severity of COVID-19, lower treatment efficiency, and higher healthcare strain. In terms of absolute count, older population in low income countries is larger than that in high income countries, and the prevalence of risk factors such as lack of hygiene facilities, handwashing soap, and water are greater (Schellekens and Sourrouille 2020). Higher viral load has been suggested to be linked to more severe disease (Zheng et al. 2020). Healthcare resources are usually low in low-income countries (Fryatt et al. 2010). Even before the pandemic, developing countries particularly had challenges to collect, verify, and aggregate reliable data in timely manner due to lack of resource, communication and technological development (Bram et al. 2015). And the pandemic might have accentuated the pre-existing challenges (Schellekens and Sourrouille 2020). The extent of bias is difficult to know, including whether it is still in early stage of infection in developing countries. Therefore, other possibilities, such as data quality and the stage of infection spread may need to be considered in interpreting the results and need further investigation in the future.

### Proportion of smokers and COVID-19 CFR

The proportion of female smokers was positively associated with COVID-19 CFR in the overall analysis, but the association diminished when the analysis was stratified by economical level of the countries/territories. This is counterintuitive given that severer COVID-19 was associated with male sex due to possibly immune system and hormone levels (Channappanavar et al. 2017) and smoking (Cai 2020; Vardavas and Nikitara 2020). The observed association between smoking in females and COVID-19 CFR might be due to that the proportion of female smoker reflected broadly income level of a country (Figure 6a). Linking to the theory of diffusion of innovation, the spread of smoking habit has been illustrated to take several stages of rise, leveling and decline, from rich to poor, men to women, and young to old (Huisman et al. 2005; Khlat et al. 2016; Pampel 2006). In the early phrase, the prevalence of smoking increase in men, and women take up smoking about a few decades later. Subsequently, male smoking start to decline and female follows later on. This pattern was found to spread from rich to poor countries. In general, Asian and African countries tended to have low female smoking but high male smoking, while in European countries prevalence was similar between men and women (Gallus et al. 2020; Hitchman and Fong 2011). Therefore, female smoking in the overall analysis was a marker of the development of a country and it diminished when the analysis was stratified by it.

**Figure 6.**
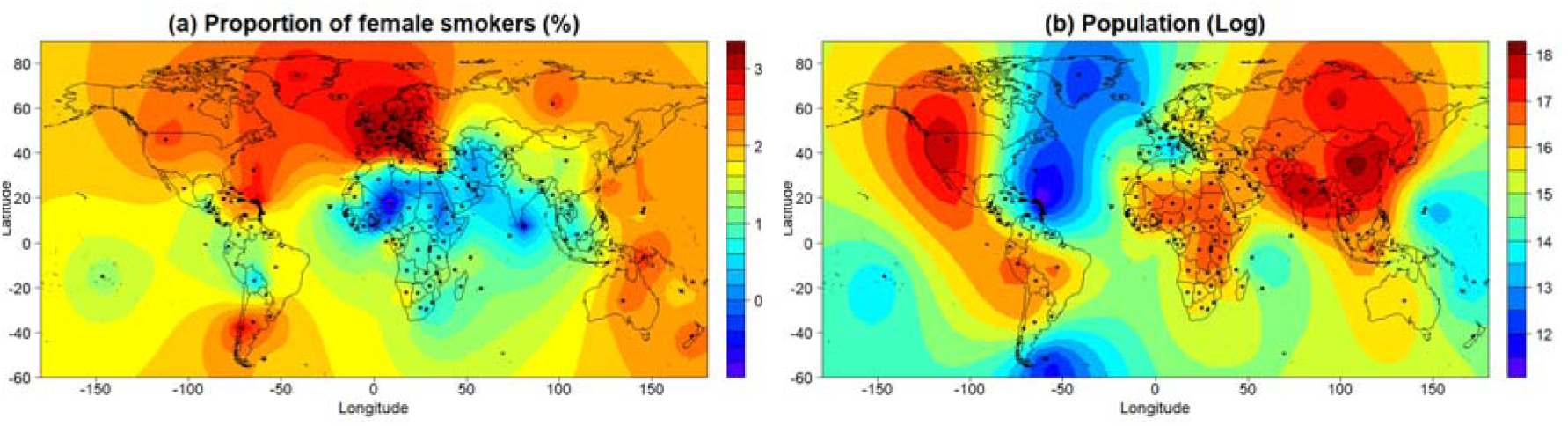
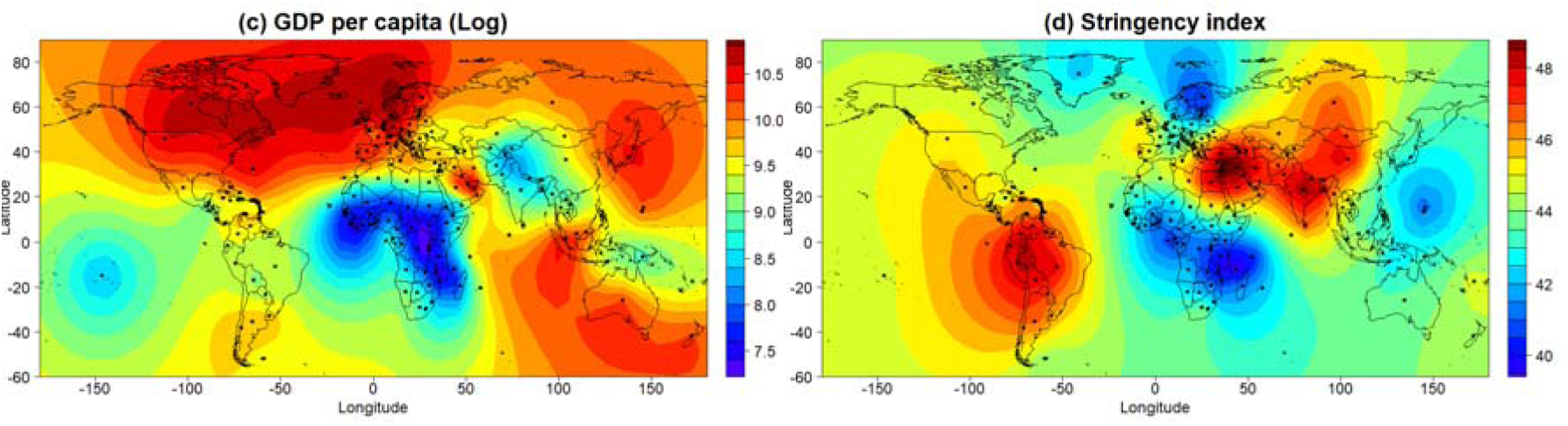
Contour plot of projected (a) proportion of female smoker, (b) population, (c) GDP per capita, and (d) stringency index

### Stringency of measures against the COVID-19 epidemic and COVID-19 CFR

According to the currently available data from the Oxford COVID-19 Government Response Tracker (Hale et al. 2020), South America and Asian took the strictest measures (Table 4 and Figure 6d), and they also had relatively lower COVID-19 CFR (Table 4). However, in our multivariable analysis controlled for other variables and spatial dependence, we did not observe a statistically significant association between stringency index and COVID-19 CRF. In contrast, stricter measures were even found being associated with higher CFR in upper-middle-income and high-income countries with active testing policies (Table 3), which seems supporting the current argument that lockdown measures might result in excess deaths in underprivileged populations and those in need are hit harder by the crisis (Melnick and Ioannidis 2020). So far, the evidence that stricter response reduced healthcare strain or treatment efficiency reflected by COVID-19 CFR globally is lacking. However, the findings need to be further examined by comparing the all-cause mortalities in previous years. Meanwhile, the reliability of the stringency index also needs to be further investigated. The relationship between socioeconomic measures against the pandemic and COVID-19 CFR is a complicated issue, which needs deeper spatiotemporal analysis with more detailed and reliable information in the future.

**Table 4.** Stringency of measures against COVID-19 epidemic and COVID-19 CRF by continent

| Continent | Stringency index |  | CRF (%) |  |
| --- | --- | --- | --- | --- |
|  | Mean | Median | Mean | Median |
| Africa | 41.57 | 43.55 | 2.33 | 1.76 |
| Asia | 46.79 | 48.85 | 2.10 | 1.24 |
| Europe | 42.57 | 43.70 | 5.15 | 4.25 |
| North America | 45.71 | 46.42 | 4.28 | 3.10 |
| Oceania | 42.62 | 40.85 | 1.47 | 0.66 |
| South America | 49.35 | 51.28 | 2.96 | 2.88 |

Noticeably, we also observed negative associations between COVID-19 CFR and CVD death rate and diabetes prevalence in some analysis, which might be partially explained by the competing risk between the deaths and/or comorbidities (Berry et al. 2010), because most of COVID-19 deaths are elderly and have one or more comorbidities (Garnier-Crussard et al. 2020; Richardson et al. 2020; Yang et al. 2020). Therefore, the COVID-19 CFR worldwide deserves deeper investigation with more detailed comorbidity information.

### Strengths and Limitations

To our knowledge, this is the first study that investigated relationship between COVID-19 CFR and demographic and socioeconomic factors globally. The study included in total 10,445,656 confirmed COVID-19 cases and 511,030 deaths worldwide. Although numerous studies have investigated the aforementioned factors related to the COVID-19 CFR, either they used much smaller sample size and investigated the question locally, or they did not approach this issue from a geospatial perspective. Our study may inspire new reflections from the healthcare workers to work together against the COVID-19 pandemic geographically and globally. International comparison of CFR may be challenging when the ascertainment of COVID-19 cases differed by country. To tackle this, we performed a sensitivity analysis using CDR. Although some risk factors, such as CVD and diabetes, showed different pattern of association, population showed consistent and positive association (see Appendix).

There are some limitations in our study. Firstly, the case-fatality analyzed here was based on the reported COVID-19 cases and deaths by countries/territories. According to the recent estimations, asymptomatic carriers and victims of COVID-19 could be as high as 10-80% in a population (Anastassopoulou et al. 2020; Day 2020; Kimball et al. 2020; Mizumoto et al. 2020; Nishiura et al. 2020; World Health Organization 2020b). However, this fraction was not taken into account in our analysis. Therefore, the case-fatality rates presented in the study might be significantly higher than the real ones. Secondly, age structure of population influences both prevalence and mortality of COVID-19, although we adjusted our analysis using the proportion of age over 60 years in populations, residual confounding largely remains. Thirdly, no detailed comorbidity information from the COVID-19 cases available in current study, which might bias the association towards an unknown direction. Finally, during an ongoing pandemic, delayed reporting occurs for both the number of cases and deaths, and strategies against the crisis also change time by time. Although we did the analysis two times using the data obtained on 17 June 17 and on July 2, and produced the same results, which suggests the bias due to delayed report might be negligible, the dynamic of the problem need to be addressed incorporating with temporal statistics methods.

## Conclusions

The average of country/territory-specific COVID-19 CFR is about 2-3% worldwide, which is higher than previously reported 0.7-1.3% and possibly due to the unreported asymptomatic cases. The COVID-19 CFR is statistically significantly associated with population size, especially in middle-income and high-income countries, which may indicate the healthcare strain and/or lower treatment efficiency in the countries with large populations. No statistically significant findings were found in low-income countries, which might be due to the challenges in data collection, communication, and verification in the countries and need to be further investigated in follow-up studies. To make global joint strategy and/or policy against the COVID-19 pandemic, spatial dependence and temporal trends must be consider in data analysis and decision making.

## Materials and methods

### Data on COVID-19 by Our World in Data

The COVID-19 dataset used in the study was downloaded from the Our World in Data website on July 2, 2020, which is a collection of the COVID-19 data maintained by the organization Our World in Data and updated daily. The dataset includes country level daily data on confirmed cases, deaths, and testing, as well as other variables of potential interest (Our World in Data 2020a; Roser et al. 2020). The data sources of the dataset including the European Centre for Disease Prevention and Control, the International Organization for Standardization (ISO), national government reports, the Department of Economic and Social Affairs of the United Nations (UN), UN Population Division, UN Statistics Division, Oxford COVID-19 Government Response Tracker, the World Bank, the Global Burden of Disease Collaborative Network, and Eurostat of the Organization for Economic Cooperation and Development (Our World in Data 2020a). There are in total 34 indices from 209 countries and territories in the dataset by July 2, 2020. The dataset was linked to the global geospatial vector database using the ISO 3166-1 alpha-2 codes for the spatial modelling (International Organization for Standardization (ISO) 2006).

### Case-fatality rate (CFR)

CFR of COVID-19 was calculated as the number of total deaths due to COVID-19 divided by the number of total confirmed COVID-19 cases by July 2, 2020, multiplied by 100. CFR was investigated in our study because it may reflect disease severity as well as efficiency of treatment and healthcare response and strain. CRF is not constant. It can vary between populations and over time, depending on the interplay between the causative agent of disease, the host, and the environment as well as available treatments and quality of patient care. For example, it can increase if the health care system is overwhelmed by the sudden increase of cases (Harrington 2020).

We also calculated crude cause-specific death rate (CDR) of COVID-19 in a sensitivity analysis and compared it with CFR. The CDR was calculated as the number of total deaths due to Covid-19 divided by the production of population and months of the data collected, multiplied by 1,000,000 (Centers for Disease Control Prevention 2012).

### Statistical analysis

The numbers of confirmed cases, tests, and tests per thousand people were not included in the analysis because they are much depending on the population, detection policy, and quarantine and isolation policy of the countries and territories. Instead, we included the stringency index in the analysis, which is a composite measure based on 9 response indicators including school closures, workplace closures, testing policy, and travel bans etc., rescaled to a value from 0 to 100 (100 = strictest response) (Haleet al. 2020). The stringency index data were obtained from the World Intellectual Property Organization website on July 1, 2020 (World Intellectual Property Organization 2020). Because the variable “proportion of the population with basic handwashing facilities on premises” has missing values in more than 50% of the countries/territories, it was also excluded from the analysis.

A subcomponent of the stringency index is the government policy on the access to COVID-19 test by 4 groups: 0, no policy; 1, only those who were both symptomatic and met specific criteria; 2, anyone symptomatic; 3, and open public testing, such as drive through testing (Haleet al. 2020). The testing policy indicator was used for stratification of the analysis.

Collinearity and multiple collinearity between the variables were examined using the Pearson’s correlation coefficient and multiple correlation coefficient, respectively (Mason and Perreault 1991). Spatial autocorrelation (or spatial dependence) is defined as the relationship between spatial proximity among some observational units and similarity among their values; positive spatial autocorrelation refers to situations in which the nearer the observational units, the more similar their values (and vice versa for its negative counterpart) (Lee 2017). This feature violates the assumption of independence among observations upon which many regression analyses are applied. Spatial autocorrelation among the fatality rates of the countries/territories was examined using a multivariate linear regression model and the Moran’s I test (Ren et al. 2014). The autocorrelation was visualized using the Matern correlation coefficient (Matérn 2013).

The Matern correlation model, a commonly used model for spatial correlated data, was fitted for our data to investigate the relationship between COVID-19 case-fatality and the demographic and socioeconomic variables. The latitude and longitude of the centroid of the countries/territories were used as random effects in the Matern correlation model (Ward and Gleditsch 2018).

Variables with skewed distribution were log transformed before entering the regression models. Multiple imputation method was used to handle the missing values in the data, and 10 imputed datasets were used for the regression analyses (Royston 2004).

Subgroup analysis was conducted by economic levels of the countries/territories according to the World Bank’s newest classification (World Bank 2020).

The associations of the studied variables with COVID-19 CDR (per 1,000,000 person months) of the countries/territories for December 31, 2019 to July 1, 2020 were also evaluated using the same methodology but using Poisson regression model, and the results were presented as appendix.

All the analysis were conducted in R 4.02 (the R Foundation for Statistical Computing, Vienna, Austria) using the package *spaMM* (Rousset and Ferdy 2014) and in Python 3.6 (Python Software Foundation) (van Rossum 1995) using the packages *geopandas* and *geoplot* (Jordahl et al. 2019).

## Data Availability

All data used in this study are publicly available and are referenced in the manuscript.

https://ourworldindata.org/

## Competing interests

The authors declare no competing interests.

## Author contributions

YC initially designed the study. All authors refined the study design and developed the concept. YC collected and analyzed the data, and drafted the manuscript with AH. All authors interpreted data and critically revised the manuscript. All authors have approved the final version for publication.

## Data sharing

All data used in this study are publicly available and are referenced in the manuscript. The R code used to run the analyses is available on request.

## Supplementary files

- GATHER statement checklist

## Appendix

**Figure S1.**
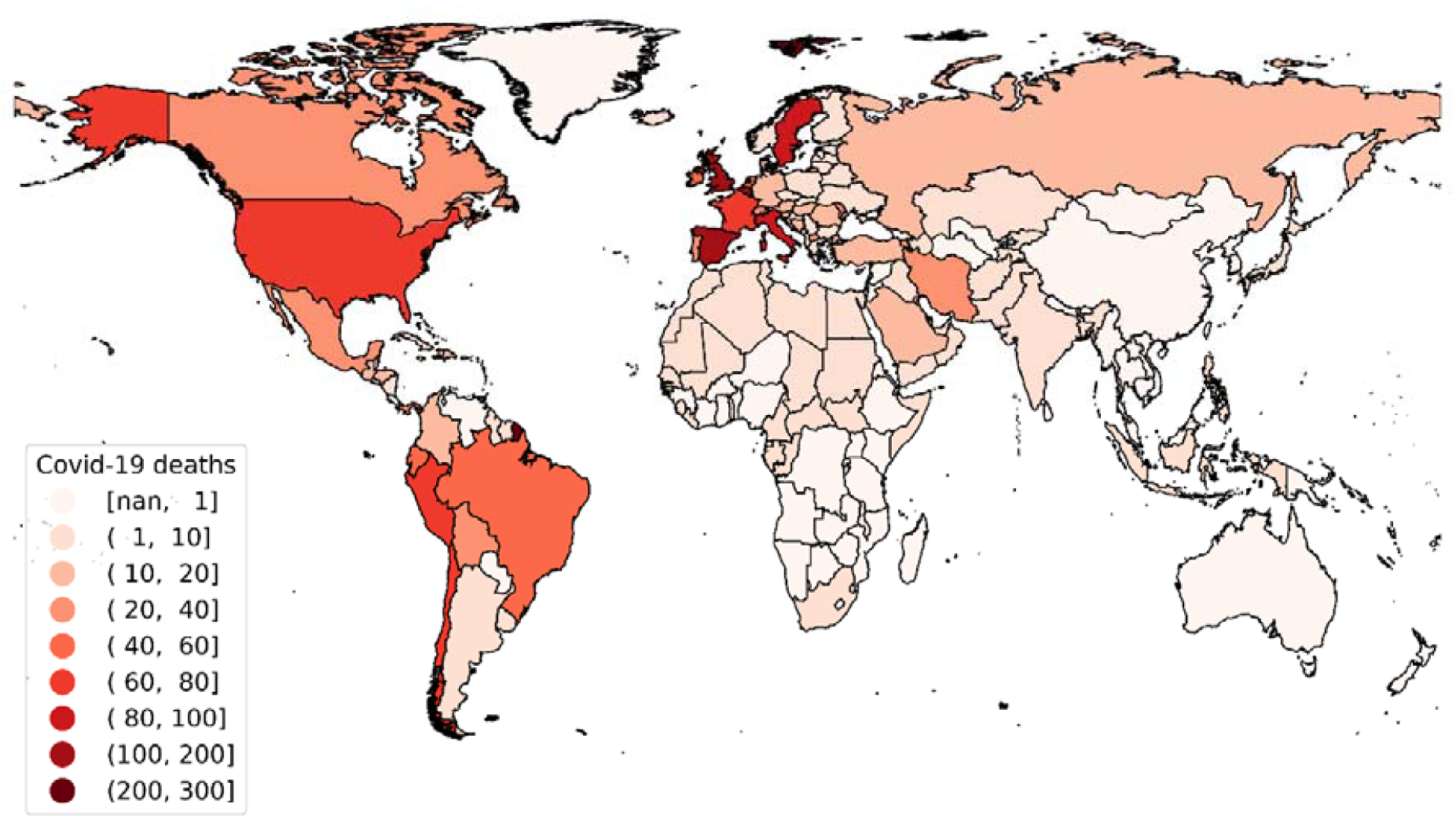
Global COVID-19 deaths (per 1,000,000 person months)

**Figure S2.**
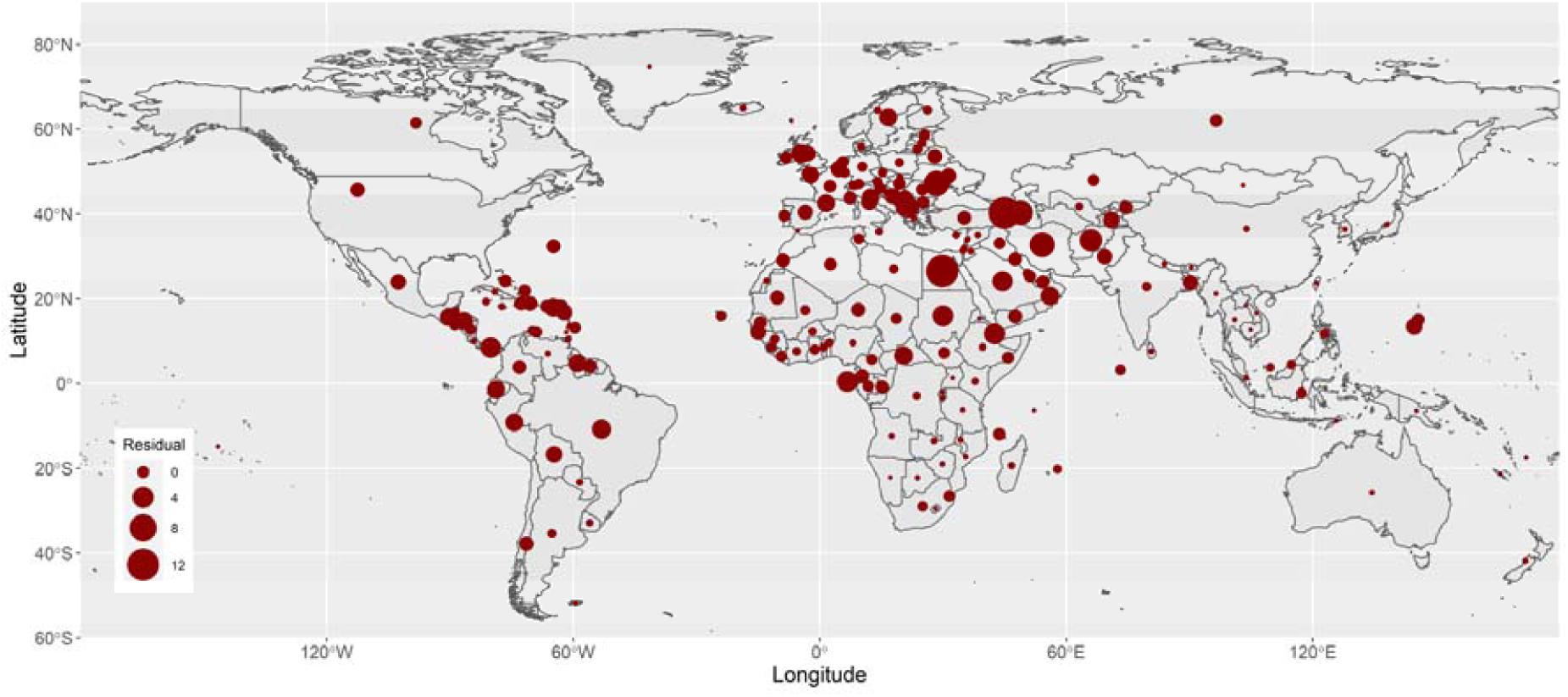
Residuals of COVID-19 deaths per 1,000,000 person months based on the common (non-spatial) multivariate Poisson regression The P-value from the Moran’s I test for the spatial autocorrelation of the residuals < 0.001.

**Figure S3.**
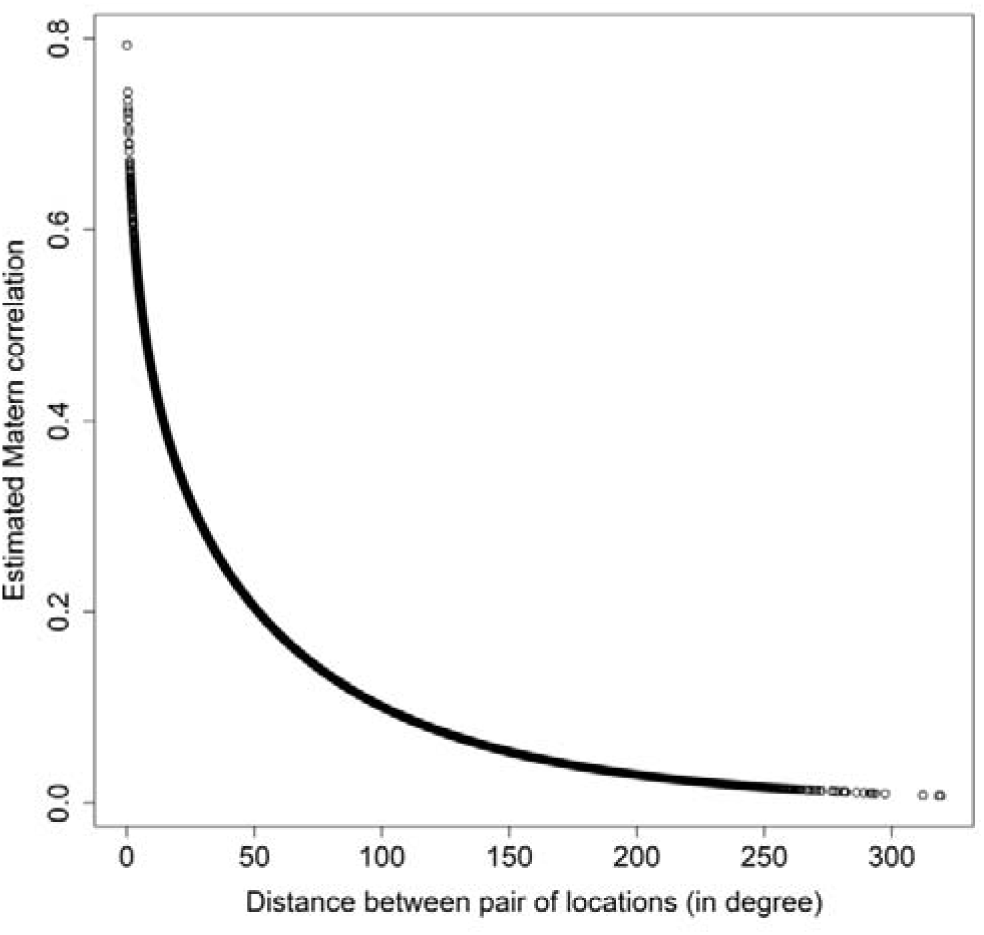
Strength and decay of the spatial autocorrelation between pair of locations (COVID-19 deaths per 1,000,000 person months) The estimated spatial autocorrelation coefficient of COVID-19 case-fatality rates between two locations against their distance is shown in Figure S3, with a strength parameter *ν* of 0.11 and a decay parameter *ρ* of 0.01.

**Table S1.** Estimation for variables’ incidence rate ratio (IRR) for COVID-19 CDR

| Variables | Unadjusted |  | Adjusted |  |
| --- | --- | --- | --- | --- |
|  | IRR (95% CI) | P-value | IRR (95% CI) | P-value |
| *Overall* |  |  |  |  |
| GDP per capita (log) | 1.69 (1.32, 2.16) | 0.000 | 1.29 (0.93, 1.78) | 0.186 |
| Population (log) | 1.10 (1.00, 1.22) | 0.054 | 1.12 (1.02, 1.23) | 0.023 |
| Population density (log) | 1.13 (0.96, 1.33) | 0.202 | 1.08 (0.92, 1.27) | 0.416 |
| Stringency index | 1.01 (0.99, 1.03) | 0.475 | 1.01 (0.98, 1.03) | 0.55 |
| Proportion of female smokers (log) | 1.31 (1.08, 1.59) | 0.017 | 1.18 (0.96, 1.44) | 0.162 |
| Proportion of male smokers | 0.99 (0.97, 1.01) | 0.391 | 1 (0.98, 1.02) | 0.587 |
| CVD death rate (log) | 0.24 (0.15, 0.41) | 0.000 | 0.41 (0.2, 0.84) | 0.046 |
| Diabetes prevalence (log) | 1.08 (0.64, 1.84) | 0.520 | 1.08 (0.63, 1.86) | 0.598 |
| Testing policy |  |  |  |  |
| 0 (Ref) |  |  |  |  |
| 1 | 0.63 (0.37, 1.08) | 0.116 | 0.59 (0.36, 0.97) | 0.058 |
| 2 | 0.79 (0.43, 1.44) | 0.438 | 0.66 (0.37, 1.18) | 0.214 |
| 3 | 1.15 (0.45, 2.9) | 0.566 | 0.74 (0.32, 1.74) | 0.551 |
| *Low-income countries* |  |  |  |  |
| GDP per capita (log) | 0.54 (0.18, 1.61) | 0.275 | 1.26 (0.41, 3.9) | 0.711 |
| Population (log) | 0.69 (0.42, 1.12) | 0.136 | 0.69 (0.43, 1.11) | 0.134 |
| Population density (log) | 0.54 (0.35, 0.85) | 0.017 | 0.82 (0.53, 1.29) | 0.411 |
| Stringency index | 0.97 (0.93, 1.02) | 0.265 | 0.99 (0.95, 1.04) | 0.807 |
| Proportion of female smokers (log) | 1.61 (1.04, 2.48) | 0.051 | 1.2 (0.67, 2.14) | 0.469 |
| Proportion of male smokers | 1.02 (0.97, 1.07) | 0.431 | 0.98 (0.91, 1.06) | 0.502 |
| CVD death rate (log) | 16.09 (4.16, 62.28) | 0.00 | 11.54 (1.73, 76.93) | 0.014 |
| Diabetes prevalence (log) | 1.56 (0.58, 4.17) | 0.392 | 0.89 (0.34, 2.35) | 0.649 |
| Testing policy |  |  |  |  |
| 0 (Ref) |  |  |  |  |
| 1 | 0.55 (0.24, 1.28) | 0.183 | 0.71 (0.26, 1.95) | 0.542 |
| 2 | 0.41 (0.09, 1.82) | 0.288 | 0.61 (0.09, 3.95) | 0.626 |
| 3 | 0.00 (0.00, inf) | 0.990 | 0.00 (0.00, 0.00) | 0.990 |
| *Lower-middle-income countries* |  |  |  |  |
| GDP per capita (log) | 0.9 (0.33, 2.47) | 0.832 | 1.5 (0.57, 3.93) | 0.461 |
| Population (log) | 0.85 (0.67, 1.07) | 0.16 | 0.93 (0.72, 1.21) | 0.63 |
| Population density (log) | 1.3 (0.88, 1.93) | 0.197 | 1.25 (0.88, 1.79) | 0.246 |
| Stringency index | 1.04 (0.99, 1.09) | 0.094 | 1.03 (0.99, 1.09) | 0.223 |
| Proportion of female smokers (log) | 1.45 (0.92, 2.28) | 0.172 | 1.04 (0.69, 1.57) | 0.633 |
| Proportion of male smokers | 0.99 (0.96, 1.02) | 0.537 | 0.99 (0.96, 1.02) | 0.367 |
| CVD death rate (log) | 0.56 (0.11, 2.75) | 0.476 | 0.58 (0.14, 2.48) | 0.485 |
| Diabetes prevalence (log) | 1.32 (0.52, 3.36) | 0.573 | 1.93 (0.86, 4.31) | 0.165 |
| Testing policy |  |  |  |  |
| 0 (Ref) |  |  |  |  |
| 1 | 0.3 (0.12, 0.76) | 0.027 | 0.32 (0.11, 0.93) | 0.080 |
| 2 | 0.21 (0.07, 0.66) | 0.016 | 0.17 (0.05, 0.61) | 0.019 |
| 3 | 0.17 (0.03, 1.17) | 0.076 | 0.08 (0.01, 0.75) | 0.048 |
| *Upper-middle-income countries* |  |  |  |  |
| GDP per capita (log) | 2.25 (0.68, 7.49) | 0.187 | 1.09 (0.34, 3.55) | 0.579 |
| Population (log) | 1.44 (1.15, 1.8) | 0.002 | 1.38 (1.03, 1.85) | 0.046 |
| Population density (log) | 0.95 (0.67, 1.34) | 0.749 | 0.94 (0.66, 1.33) | 0.712 |
| Stringency index | 0.99 (0.95, 1.04) | 0.676 | 0.98 (0.93, 1.02) | 0.473 |
| Proportion of female smokers (log) | 1.09 (0.76, 1.57) | 0.567 | 1.2 (0.86, 1.69) | 0.295 |
| Proportion of male smokers | 0.97 (0.93, 1) | 0.126 | 0.99 (0.95, 1.03) | 0.58 |
| CVD death rate (log) | 0.25 (0.08, 0.84) | 0.031 | 0.3 (0.09, 0.96) | 0.077 |
| Diabetes prevalence (log) | 0.48 (0.13, 1.86) | 0.305 | 0.77 (0.22, 2.72) | 0.704 |
| Testing policy |  |  |  |  |
| 0 (Ref) |  |  |  |  |
| 1 | 0.52 (0.21, 1.28) | 0.230 | 0.57 (0.24, 1.34) | 0.302 |
| 2 | 0.77 (0.26, 2.27) | 0.450 | 0.74 (0.26, 2.09) | 0.407 |
| 3 | 0.74 (0.06, 9.27) | 0.816 | 0.49 (0.03, 7.19) | 0.518 |
| *High-income countries* |  |  |  |  |
| GDP per capita (log) | 2.49 (0.98, 6.37) | 0.126 | 2.52 (0.96, 6.59) | 0.122 |
| Population (log) | 1.22 (1.07, 1.4) | 0.004 | 1.19 (1.02, 1.39) | 0.051 |
| Population density (log) | 1.11 (0.89, 1.4) | 0.396 | 1.14 (0.89, 1.47) | 0.353 |
| Stringency index | 1.03 (0.98, 1.08) | 0.287 | 1.02 (0.96, 1.07) | 0.552 |
| Proportion of female smokers (log) | 1.19 (0.8, 1.77) | 0.372 | 0.97 (0.54, 1.74) | 0.406 |
| Proportion of male smokers | 1 (0.96, 1.04) | 0.596 | 1.00 (0.95, 1.05) | 0.433 |
| CVD death rate (log) | 0.22 (0.09, 0.55) | 0.006 | 0.5 (0.16, 1.6) | 0.413 |
| Diabetes prevalence (log) | 0.58 (0.23, 1.47) | 0.298 | 0.69 (0.21, 2.31) | 0.567 |
| Testing policy |  |  | 2.52 (0.96, 6.59) | 0.122 |
| 0 (Ref) |  |  |  |  |
| 1 | 0.81 (0.29, 2.28) | 0.468 | 0.60 (0.24, 1.54) | 0.449 |
| 2 | 0.67 (0.23, 1.97) | 0.437 | 0.66 (0.25, 1.75) | 0.376 |
| 3 | 1.08 (0.27, 4.3) | 0.611 | 1.01 (0.28, 3.69) | 0.619 |

GDP, gross domestic product; CVD, cardiovascular disease; CI, confidence interval.

**Figure S4.**
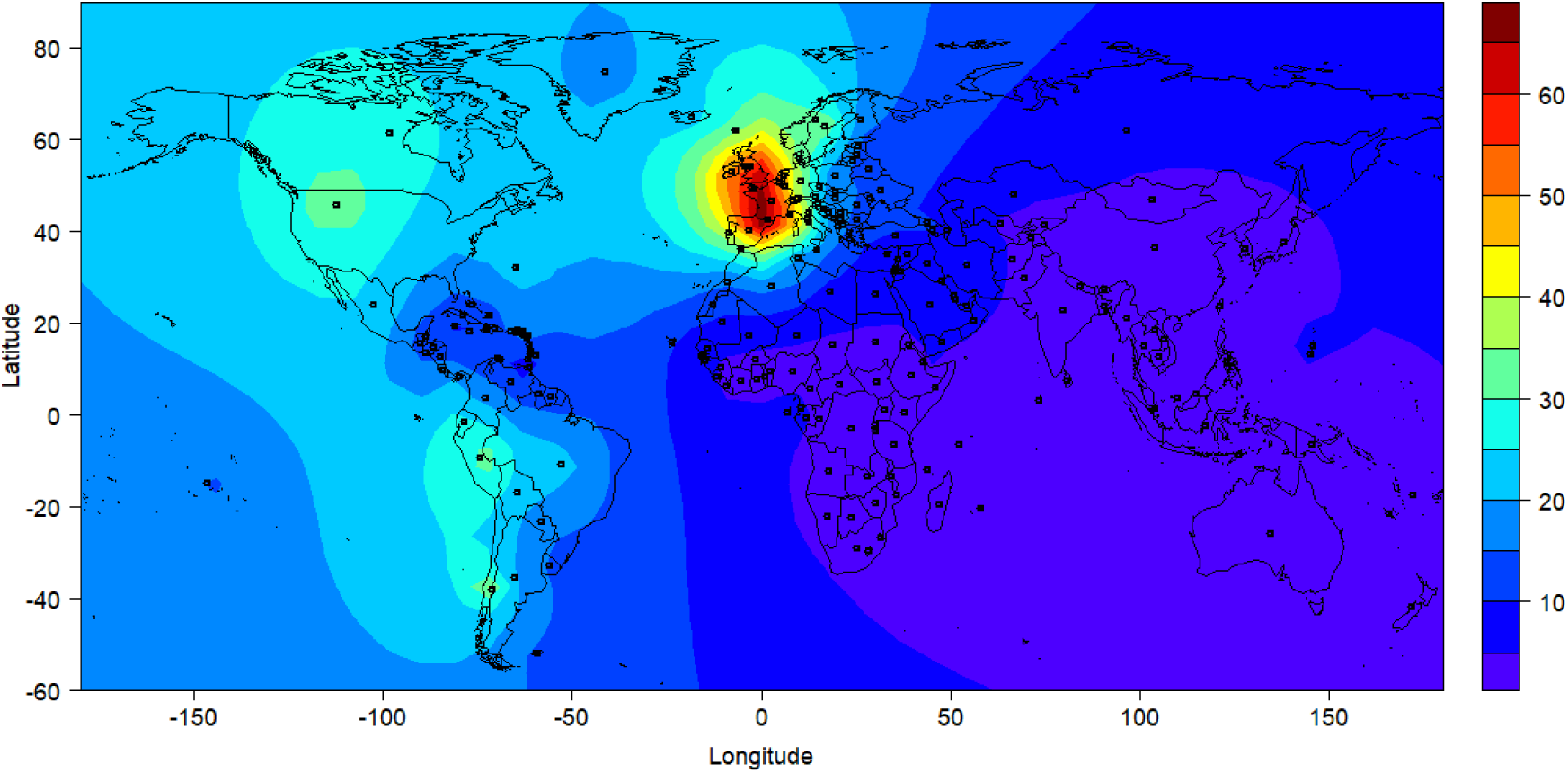
Contour plot of estimated COVID-19 deaths (per 1,000,000 person months)

**Table S2.**
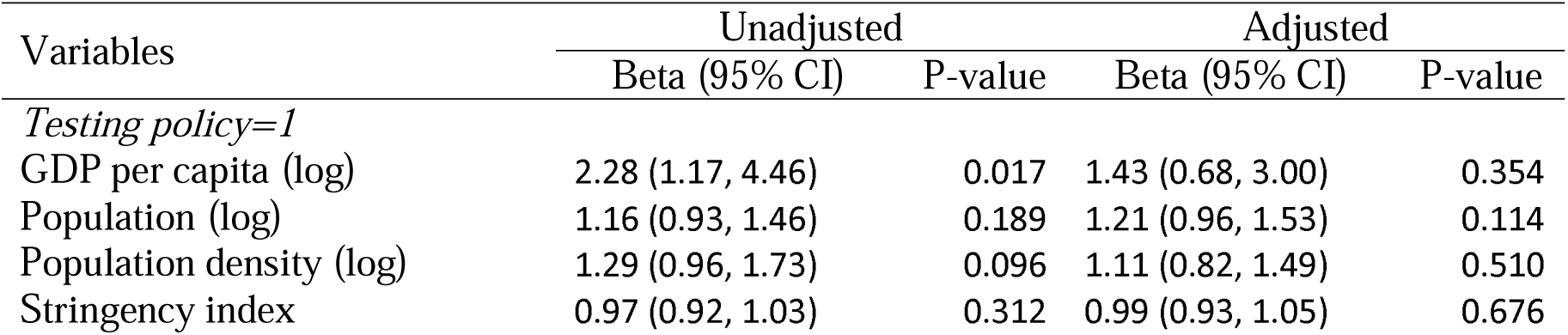
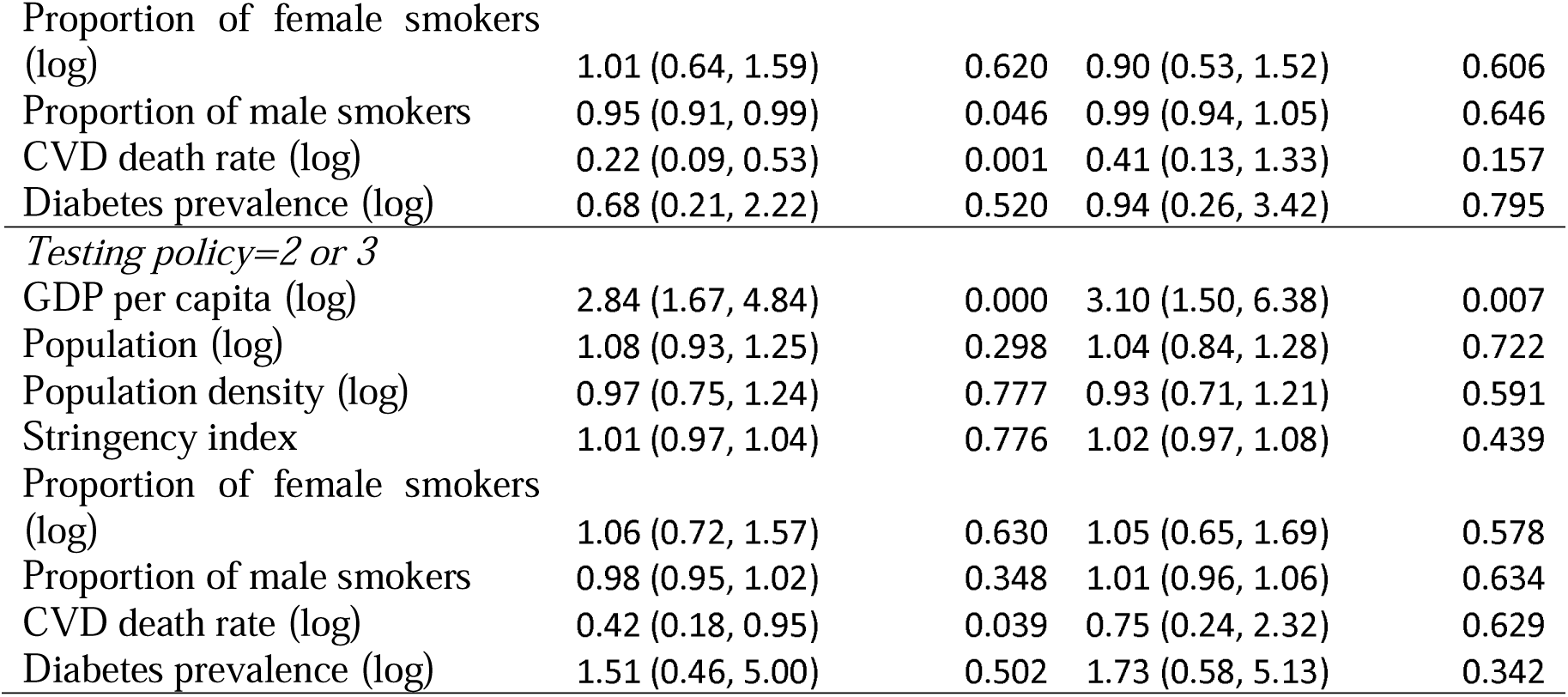
Estimation for variables’ incidence rate ratio (IRR) for COVID-19 CDR by testing policy in upper-middle-income and high-income countries

## Notes

### Competing Interest Statement

The authors have declared no competing interest.

### Funding Statement

No funding for this study

### Author Declarations

The data used in the study are freely available in the open source for research. There are no individual data available in the dataset. The owner of the data gives the permission to use, distribute, and reproduce the data in any medium. Therefore, no private and confidential information could be disclosed in the study, and ethical approval is not applicable.

